# Feasibility of an online Langerian mindfulness program for stroke survivors and caregivers

**DOI:** 10.1101/2021.08.05.21261674

**Authors:** Marika Demers, Francesco Pagnini, Deborah Phillips, Brianna Chang, Carolee Winstein, Ellen Langer

## Abstract

**Background:** Mindfulness is promising for individuals with neurological disorders and their caregivers to improve psychological well-being. The potential application of a Langerian mindfulness intervention, focused on attention to variability, however, is still unknown.

**Objective:** To determine the feasibility (usability, satisfaction, and potential effectiveness on psychological well-being) of an online mindfulness intervention for stroke survivors and caregivers.

**Methodology:** Using mixed-methods, 11 stroke survivors and 3 caregivers participated in a 3-week, online, Langerian mindfulness intervention. A semi-structured interview assessed the intervention’s usability and gathered feedback. Self-reported measures about psychological well-being were documented remotely three times (pre, post, one-month follow-up).

**Results:** Qualitatively, participants were highly satisfied with the intervention, reported subjective benefits, but the usability of the online platform was poor. None of the self-reported measures changed over time.

**Conclusion:** This study provided evidence of feasibility of an online Langerian mindfulness intervention in a new population: stroke survivors and caregivers.

## Introduction

Stroke is a heterogeneous condition and a leading cause of disability (Mayo et al., 2002). Stroke survivors have poor perceived physical health (Patel et al., 2006), high incidence of anxiety and depression (Ayerbe et al., 2013; Campbell Burton et al., 2013), and high levels of dissatisfaction in life, which significantly impact activity and participation (Hartman-Maeir et al., 2007). The psychosocial impact of stroke extends to loved ones who serve in a caregiver role (Rigby et al., 2009). Caregivers are crucial in preserving rehabilitation gains and independence levels, maintaining community participation, and conserving the long-term well-being of stroke survivors (Rigby et al., 2009; Visser-Meily et al., 2006). The proportion of caregivers experiencing significant burden is estimated to range between 25-54% (Rigby et al., 2009). Compared to age-matched peers, caregivers have diminished general health and vitality, and higher prevalence of depression (Morimoto et al., 2003).

Currently, some of the most promising clinical treatments for distress reduction and psychological well-being improvement are based on the concept of *mindfulness* (Pagnini & Philips, 2015). Mindfulness is multifaceted, but here, it is defined using Ellen Langer’s non-meditative approach as the process of actively making new distinctions about a situation and its environment, or its *current context*, rather than relying on previous categorizations. Previous studies indicate that many presumed physical or psychological limits result from mindlessness, and participants may benefit from increasing mindfulness (Langer, 2012). While investigation into clinical applications of mindfulness has been initiated, there is still significant clinical potential to explore the mindfulness construct as it relates to chronic disease.

Mindfulness interventions based on Langer’s mindfulness theory were shown to increase mindfulness in the general population and other cohorts (Alexander et al., 1989; Grant et al., 2004; Langer, 2000, 2009). One example is an online Langerian mindfulness intervention for individuals with amyotrophic lateral sclerosis (ALS) and their caregivers. The mindfulness group reported higher quality of life and lower levels of negative emotions, anxiety, and depressive symptoms, and for caregivers, lower levels of care burden compared to a waitlist control (Pagnini et al., 2021). Most differences remained stable over time, indicating a sustained effect of the mindfulness intervention, at least short-term. In stroke survivors, three systematic reviews reported a positive trend for mindfulness-based interventions on psychological, physiological, and psychosocial outcomes. However, stronger evidence remains to be established with robust clinical trials (Abbott et al., 2014; Lawrence et al., 2013; Ulrichsen et al., 2016). Given the favorable results for individuals with neurological conditions and their caregivers, a stroke-specific online mindfulness intervention is promising to increase mindfulness, decrease anxiety and depression, and change patients’ beliefs about their disability. Most mindfulness-based interventions use meditation and contemplative practices to promote mindfulness (Pagnini & Philips, 2015). These require relatively significant time and effort, limiting potential real-life application, particularly in clinical contexts and with caregivers (Jani et al., 2018). The Langerian approach focuses on attention to variability and provides easily accessible cognitive exercises inducing openness, cognitive flexibility, and creativity (Pagnini et al., 2016) (see Table 1 for definitions and examples).

**Table 1.** Structure of the mindfulness program

| Topic | Definition | Example |
| --- | --- | --- |
| Attention to variability | The key concept is that the world around us, and each one of us, is always changing. If we can notice subtle changes in the external world, as well as in how we feel, we will find that negative feelings and symptoms are not always present. Thus, you can change the usual tendency to view things as if they are unchanging – a mindless construct of the world. | Try to find 5 ways in which the weather is different today than yesterday. |
| Positive and negative events | The same event may be seen as *either* positive or negative, depending on one's perspective. If we change our point of view, then our perception about what we were certain is a negative event can, from that different perspective, be viewed as positive. That is, we can reframe a negative perception of an event by answering a simple question: "In what ways might this be good for me? Or "What about this situation could be seen as good?" After a stroke, we can try to find a different perspective about this condition. We see that our experience of this condition reflects the view we take rather than of the condition itself. | You miss a bus to meet an important person in your life. Try to find 5 reasons how that could be positive. |
| Unpredictability | A mindful view of the future acknowledges its ultimate unpredictability, as is the course of the one's disability. Such unpredictability opens possibilities for a more positive perception of the future than had been conceived. | Can you predict the course of stroke recovery? If so, please find 3 ways in which your certainty could be faulty. For each of those reasons, describe the way in which it is both positive and negative. |
| Sense Making | Disability and symptoms tend to be perceived as something that stays unchanging. However, it is unlikely that all symptoms are always present at the same intensity, or the impairments stay the same. Reflecting about symptoms and ability to do meaningful things allows for understanding their variability. Making sense of this can help develop an understanding that there are often many | Think about your past, particularly the most meaningful moments in your life. Can you see how these events may have led to one another or somehow |
|  | reasons for their variability and feelings of increased control and awareness follow. | be connected in some way? |
| Novelty seeking and novelty producing | The attitude of paying attention to new elements requires an open and curious orientation toward the world and towards oneself. Novelty producing actively creates new categories of thinking rather than relying on previously constructed categories and distinctions about the world. Novelty seeking relates to the concept of flexibility or using feedback from the environment to make any responsive adaptations to one's thinking and behavior. | We invite you to consider this: What are 5 ways you could make something in your home more personal and appealing to you? |

Our research team adapted the mindfulness intervention initially developed for individuals with ALS (Pagnini et al., 2021) with the aim to increase stroke survivors’ and caregivers’ mindfulness and improve psychological state. The Desire2Learn platform (D2L, Kitchner, Canada) was used to deliver the intervention, a platform used for higher education, but not adapted for clinical populations. There is a need for a feasibility study to build the foundations to support intervention trials and specifically 1) evaluate the acceptability and suitability of the intervention and the study procedures for stroke care, 2) test usability of the D2L platform to deliver the intervention, 3) assess the preliminary impact of the intervention on quality of life and well-being (Tickle-Degnen, 2013).

This study aims to evaluate the usability and satisfaction of a three-week online mindfulness intervention for stroke survivors and their caregivers. The secondary exploratory aim is to estimate the potential short-term impact on quality of life and psychological well-being. We hypothesized that the mindfulness intervention will be well accepted and relevant. It is anticipated that minor modifications will be suggested to improve understandability and relevance.

## Methods

### Study design

This feasibility study used a mixed-methods convergent design (Creswell & Plano-Clark, 2018). A qualitative semi-structured interview performed post-intervention allowed a deeper understanding of users’ perceptions of the mindfulness intervention. Quantitative self-reported measures were used at three time points (pre-, post- and one-month follow-up) to capture changes in psychological well-being. The mixed-methods approach allowed us to collect rich, comprehensive, and actionable data and compare qualitative and quantitative findings to guide the development of a larger intervention study.

### Participants

Community-dwelling stroke survivors and their loved ones donning the caregiver role were purposefully recruited from September 2020 to July 2021. Recruitment was from an existing IRB-approved database and advertisements through community support groups. Participants were included if they had Internet access on computer, tablet and/or smartphone. Individuals with severe language impairments, those who participated in regular meditation or in a mindfulness program in the past three months, or were employed caregivers were excluded. A sample of ten stroke survivors was targeted to allow sufficient richness of the qualitative data but was not powered for the secondary aim. This study was approved by the University of Southern California (USC) Health Sciences Campus IRB. All participants were informed of the risks and benefits, that their participation was voluntary, and provided electronic informed consent collected through REDCap (Vanderbilt University, Nashville, TN).

### Intervention

The three-week intervention consisted of daily mindfulness exercises and education, with one unique module for stroke survivors and one for caregivers. The intervention was accessible via a computer or a smartphone and was hosted on the USC Biokinesiology and Physical Therapy continuing education platform D2L. The content was adapted by experienced clinicians and researchers, stroke survivors, and mindfulness experts. The intervention focused on attention to variability (particularly attention to observed or felt variability in symptoms) as an overarching construct. Four new topics - positive and negative events, unpredictability, sense making and novelty producing - tapping into different aspects of attention to variability were introduced. The general structure of the intervention mixed cognitive exercises from the five topics. All exercises for the stroke survivors included audio recordings, but the caregiver module only included written text.

### Procedures

Initially, participants met with the first author (MD) using HIPAA compliant Zoom platform (Zoom Video Communications Inc., San Jose, CA) to review the study procedures, obtain consent, collect socio-demographic information, and complete the assessments via REDCap (see Figure S1 for study timeline). MD oriented participants on how to navigate the online platform. Each participant received a unique identification number and password to access D2L. After a demonstration, participants connected to the platform and accessed the first day’s content. MD recorded technical issues, difficulty experienced, assistance provided, and spontaneous feedback reported by participants. Participants were instructed to accomplish exercises at least five days per week. Follow-up emails were sent 48 hours after the beginning of the intervention and every week after to confirm adherence and remind participants to complete the daily exercises. Participants could contact the research team to report difficulties.

### Qualitative data collection

After the intervention, a semi-structured interview lasting up to 30 minutes was done over Zoom. The interview was video recorded and consisted of open-ended questions about feelings, beliefs, reactions to the intervention, accessibility of the D2L platform, technical issues, adherence, clarity and relevance of the content, perceived benefits, and suggestions to improve user experience (see Supplementary Material for the interview guide).

### Quantitative data collection

The exploratory outcome measures were collected remotely at 3 time points: pre-, post- and at one-month follow-up. For stroke survivors, the self-reported measures were: 1) Stroke Impact Scale (SIS) (Duncan et al., 2003), 2) Hospital Anxiety and Depression Scale (HADS) (Zigmond & Snaith, 1983), 3) Perceived Stress Scale (PSS) (Kupst et al., 2015), and 4) Single-Item Sleep Quality Scale (SQS) (Snyder et al., 2018). For caregivers, the self-reported measures were: 1) World Health Organization Quality of Life-BREF (WHOQOL-BREF) (World Health Organization, 2004), 2) HADS, 3) Zarit Burden Interview (Zarit et al., 1980), 4) PSS, and 5) SQS (see Supplementary material for a description of each measure). Participants were instructed to consider the impact of the global pandemic when answering the questions. The adapted Post-Study System Usability Questionnaire (PSSUQ) was administered post-intervention to assess D2L platform usability to host the mindfulness intervention (Lewis, 1992).

### Qualitative data analysis

A descriptive qualitative approach was used to understand feasibility (Colorafi & Evans, 2016). The semi-structured interviews were transcribed verbatim. Two research team members independently analyzed the transcripts using the Braun & Clarke (2006) six-step framework for inductive thematic analysis (see Supplementary material for detailed data analysis and reflexivity). The thematic analysis was performed separately for stroke survivors and caregivers, and overarching themes across both groups were identified. Any disagreements in coding were resolved through discussion and an audit trail of the decisions made was maintained to promote trustworthiness (Shenton, 2004).

### Quantitative data analysis

Descriptive statistics were used for stroke survivors’ and caregivers’ data. Moreover, for stroke survivors, non-parametric Friedman tests were used to examine whether the mindfulness intervention had an impact on quality of life and well-being measures, since the data were non normally distributed. All statistical procedures were done with JASP version 0.14.1.

### Integration of qualitative and quantitative data

Once the qualitative and quantitative analyses were completed, we integrated both data to better interpret the findings and draw overall conclusions on the feasibility of the intervention.

## Results

Twelve participants (ten chronic stroke survivors and two spouses acting as caregivers) completed the intervention and follow-up assessment, and one additional stroke survivor was lost at follow-up (socio-demographics summarized in Table 2). One additional participant was enrolled in the study but never started the intervention due to an unexpected change in medical status. Another participant dropped from the study after one week due to medical reasons. One additional caregiver supported her parents to complete the intervention. She participated in the interview but did not complete the assessments. Only three participants were familiar with mindfulness, including previous participation in guided meditation or mindfulness.

**Table 2.** Socio-demographics of participants

| **Characteristics** | **Stroke survivors**<br>**(n=11)** | **Caregivers**<br>**(n=3)** |
| --- | --- | --- |
| **Gender** (Female, %) | 3 (27.3%) | 3 (100%) |
| **Age** (Mean ± SD) | 65.1 ± 12.1 | 59.7 ± 21.5 |
| **Race** (n, %) |  |  |
| Asian | 3 (27.3%) | 2 (66.7%) |
| White or Caucasian | 7 (63.6%) | 1 (33.3%) |
| More than one Race | 1 (9.1%) | 0 |
| **Ethnicity** (n, %) |  |  |
| Hispanic | 1 (9.1%) | 0 |
| Non-Hispanic | 10 (90.9%) | 3 (100%) |
| **Ambulation** (without assistive device*, n, %) | 4 (36.4%) | 3 (100%) |
| **Living situation** (n, %) |  |  |
| Alone | 3 (27.3%) | 0 |
| With spouse | 4 (36.4%) | 2 (66.7%) |
| With family | 4 (36.4%) | 1 (33.3%) |
| **Time since stroke** (years, mean ± SD) | 4.7 ± 3.7 | n/a |
| **Hemisphere affected by the stroke** (Left, n, %) | 4 (36.4%) | n/a |
| **Stroke type** (Ischemic stroke, n, %) | 7 (63.6%) | n/a |

Legend: \* Walking independently without assistive device or physical assistance.

### Qualitative results

Four main themes emerged from the qualitative data analysis: 1) Satisfaction with and adherence to the mindfulness intervention, 2) Poor user experience with the web-based platform, 3) Varied perceived benefits from the intervention, and 4) Importance of tailoring the mindfulness intervention (see Supplementary material for a conceptual map and additional quotes). The themes reflected the participants’ perspectives towards the intervention, the difficulties experienced, and potential changes to implement.

#### Theme 1 – Satisfaction with and adherence to the mindfulness intervention

The first theme describes the knowledge and insights gained from participating in the mindfulness intervention. Eight participants expressed high satisfaction and qualified the intervention as ‘good’, ‘very good’, ‘enlightening,’ or ‘interesting’. The intervention aligned well with or exceeded participants’ expectations. They appreciated that mindfulness practice was accessible, easy to implement in everyday activities, and encouraged reflection. Participants liked that the exercises were simple and easy to understand. One participant found some exercises were emotionally challenging. Three additional participants had more nuanced opinions of the intervention and expressed being somewhat satisfied with the program. All reported that the exercises offered were not new to them, despite not being familiar with mindfulness. One found some exercises bothersome, as they did not consider her stage in stroke recovery and her capacity. She also reported that some of the language was negative. Another participant felt the intervention lacked direct feedback from a therapist. One felt it was more relevant early post-stroke.

S04: “*I enjoyed [the mindfulness intervention]. I got to see a little bit more targeted mindful practice, geared towards me and post-stroke recovery. So, yes, I did enjoy that*.”

C01: “*I didn’t have any expectations, but I was pleasantly surprised*.”

Participants adhered well to the intervention timeline and occasionally skipped days because they were busy or tired. Some participants maintained a consistent schedule to facilitate adherence or completed two days at once to compensate for missed days. The pandemic allowed most participants to have the time to take part in this program, as many usual activities were disrupted. One participant had many groups/interventions on the computer, which limited his willingness to do another online intervention.

S03: “*I got to confess to you, I would miss days and then I would go back, I would do a couple of days at a time*.”

C01: “*COVID has us doing not as much, so that [intervention] was helpful*.”

The number of exercises was considered appropriate and took 15-30 minutes to complete. Opinions about the ideal length of the intervention varied: some participants preferred a two-week intervention, others found the length appropriate, and one wished it lasted longer.

S03: “*It was right at the limit I would say it. I mean, if you would have gone on for another week, it would’ve been more than I wanted. It was pretty perfect*.”

#### Theme 2 - Poor user experience with the web-based platform

The second theme addresses the usability and accessibility of the D2L platform, accessed on a smartphone, tablet, or computer. The D2L platform was not perceived as user-friendly (see Table 3 for suggestions for improvements). Important issues with accessibility were identified: Features, such as tests and grades, were distracting; The content was not directly accessible after signing in; The browser did not systematically load the page where the participant left off, requiring them to go through the table of contents each time they logged in; The time to download videos was long, and consequently most participants skipped the videos. Two participants mentioned that the pictures were unnecessary and distracting. Participants liked the checkmark feature that indicated which exercises were completed. Four participants experienced technical issues with their Internet connectivity or their device, which limited their ability to follow the intervention.

**Table 3.** Suggestions for improvements to the Mindfulness program

| Items to address | Quotes | n |
| --- | --- | --- |
| Tailor exercises based on abilities, stages of recovery and living situation | S10: "So sometimes it would say things they would ask me to do things that I wasn't capable of doing. I'm not for instance going [...] I'm not going to the store right now for instance and it might say 'go look at a flower' or something that wasn't available." | 7 |
| Ensure that the exercises reflect the current COVID-19 situation | S04: "In these COVID time, no one is taking showers everyday (laughs). No, but there are things out of sync to what normally happens or when it happens." | 2 |
| Simplify access to the content | S05: "I felt I should be able to log in and be where I left off the time before and go forward. But every time, I had to log in, then find the program, then find where I was, and it wasn't intuitive the way you do it and sometimes it wouldn't go. You have to go to portal [...] like it wasn't like you click on one thing." | 4 |
| Remove the pictures and artwork | S07: "I think the only part that perhaps is unnecessary was the artwork." | 2 |
| Offer a place for participants to record their thoughts and ideas after each exercise | S01: "I think that somebody could if they had a place where they could type in some notes. [The participant] could give you a response that would maybe be more mindful about how that topic area affected them." | 2 |
| Change the wording used to be more inclusive | S11: "Sometimes I had trouble with particular articles like it said: 'come up with three ideas why this might be a good thing or a bad thing'. I could only come up with one idea." | 4 |
| Provide an overview of the daily exercises and specify times at which each exercise should be done | S04: "It would be nice to have maybe like: 'do this in the morning', 'do this in the evening', 'do this...' so that you sort of [...] I think there could be a little bit of structure around when in the time of the day some of these prompts could come up." | 3 |
| Offer audio recordings for the entire program | S11: "It would be good if [the introduction to each module] would be recorded also, because some people have a hard time reading after having had a stroke or a hard time concentrating." | 2 |
| Add a concluding statement at the end of the program | S05: "at the end of this, you will A, B, C, D. There was no feel for that. Like at the end of this, you just feel nothing. I mean, even if they said you may not notice any changes or anything, but I felt like I was supposed to know at the end of this something." | 1 |

S03: “*Accessing [the website] was [difficult] because I had a username and a password. I struggled with that a little bit. You can be one character off and it’s a problem. I’m not all that technically acclimated. I’m average I would say*.”

S01: “*The nice thing is that when you go to the content page, it shows you the check marks of what of the days you’ve completed and a couple of ‘em I did not remember doing*.”

The presence of audio-recordings for the stroke survivors’ module contributed to the user experience of the platform. Six participants used only the written text and four preferred the written text with the audio recordings. The audio recordings were noted as useful, even if unused. One caregiver wished audio recordings were available for the caregiver module.

C03: “*I wish the caregiver portion had an audio component […] My mom would have felt more competent if she could just listen to it while cleaning or whatnot*.”

S11: “*I like having the words to read and having the recording*.”

#### Theme 3 – Varied perceived benefits from the intervention

The third theme encompasses the perceived gains related to the mindfulness exercises and education. The most common ideas were that the intervention increased participants’ general knowledge about mindfulness and helped change their mindsets, attitudes, and viewpoints. Many participants gave examples of how they applied mindfulness in their daily lives. Participants highlighted that the intervention stimulated discussion with their loved ones and gave them tools to navigate challenging or stressful situations. Two participants with poor sleep quality reported that the program offered strategies to go back to sleep.

S03: “*It was, overall, very beneficial to me. Mindfulness was kind of like shifting your viewpoint just a little bit and that little bit was helpful*.”

C01: “*It’s sort of like a meditation, but active meditation. It helps you get your mind from a negative thought to a positive thought*.”

#### Theme 4- Importance of tailoring the mindfulness intervention

The fourth theme describes the strong perception that a mindfulness intervention should be personalized and adapted to individual situations, including a global pandemic. Both stroke survivors and caregivers discussed how stroke changed their lives. Stroke survivors mentioned how stroke disrupted their routine, meaningful activities, roles, and social participation. Most exercises were considered relevant. However, participants felt that some exercises were not targeted to their capacity, stage of recovery, or living situation. For example, one exercise encourages stroke survivors to use their paretic hand to perform activities, which was impossible for those with severe impairments, and consequently such exercises created frustrations. One stroke survivor mentioned that some exercises required to express emotions, which conflicted with her own culture. Perception of stroke recovery differed between participants, with high dissatisfaction with their recovery, often due to slower recovery or persisting impairments. Outlook on life changed at different time points post-stroke, which emphasized the importance of tailoring exercises.

S01: “*Well obviously, I wish I had recovered more especially on my left side and there have been some other impacts of my stroke over the years, other than the immediate ones. I’ve definitely had some physical issues that have come up in recent years that are sort of manifestations of the stroke or the impact on my brain are more noticeable now*.”

S11: “*If you’ve had a stroke, after a certain time, you’ve adjusted your life to a certain way*.”

The COVID-19 pandemic greatly impacted all participants and heightened uncertainties. Most participants expressed how the pandemic impacted their answers. Three participants mentioned that some exercises were not aligned with the global events, stressing the need for a mindfulness intervention to be flexible to individual situations and needs.

S04: “*It is hard to say that COVID didn’t influence anything. Every single aspect of life is affected by [COVID-19]*.”

### Quantitative results

The self-reported measures took between 10-90 minutes to administer remotely. Two caregiver-stroke dyads required assistance from a researcher to complete the assessments. Among stroke survivors, there were no significant changes in scores for the SIS, HADS, PSS, with high variability between participants (Table 4). The HADS showed a trend towards reduced anxiety and depression, with five participants showing lower levels of anxiety and depression post-intervention (Figure 1). Sleep quality improved from pre to post intervention for four participants, but this was not maintained at the one-month follow-up. During the qualitative interview, participants mentioned extenuating factors, such as pain or stress from the US elections, to explain sleep quality deterioration. The caregiver sample was not large enough to assess changes from the intervention, and no clear changes were noted.

**Table 4.**
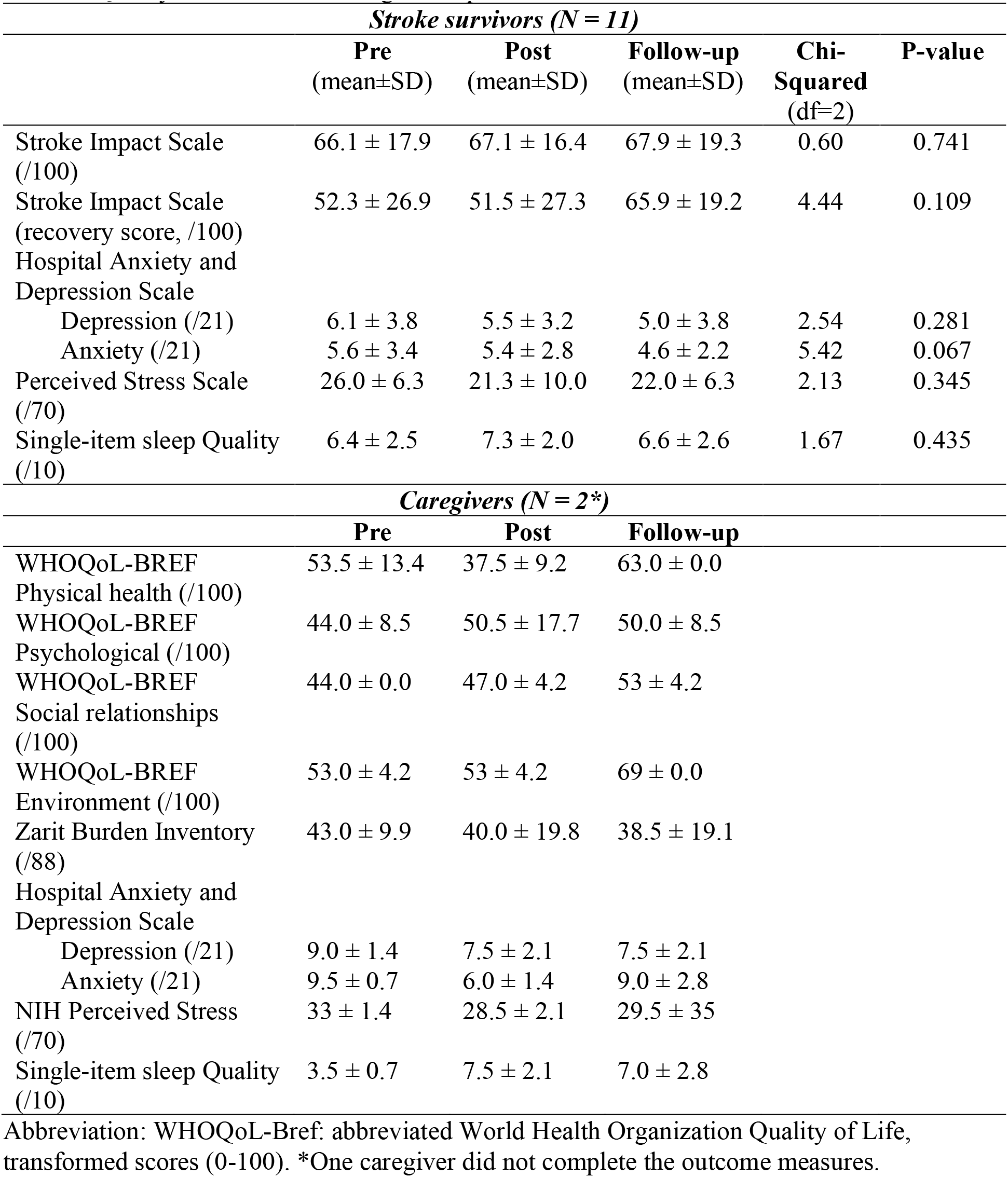
Quality of life and wellbeing self-reported measures

**Figure 1.**
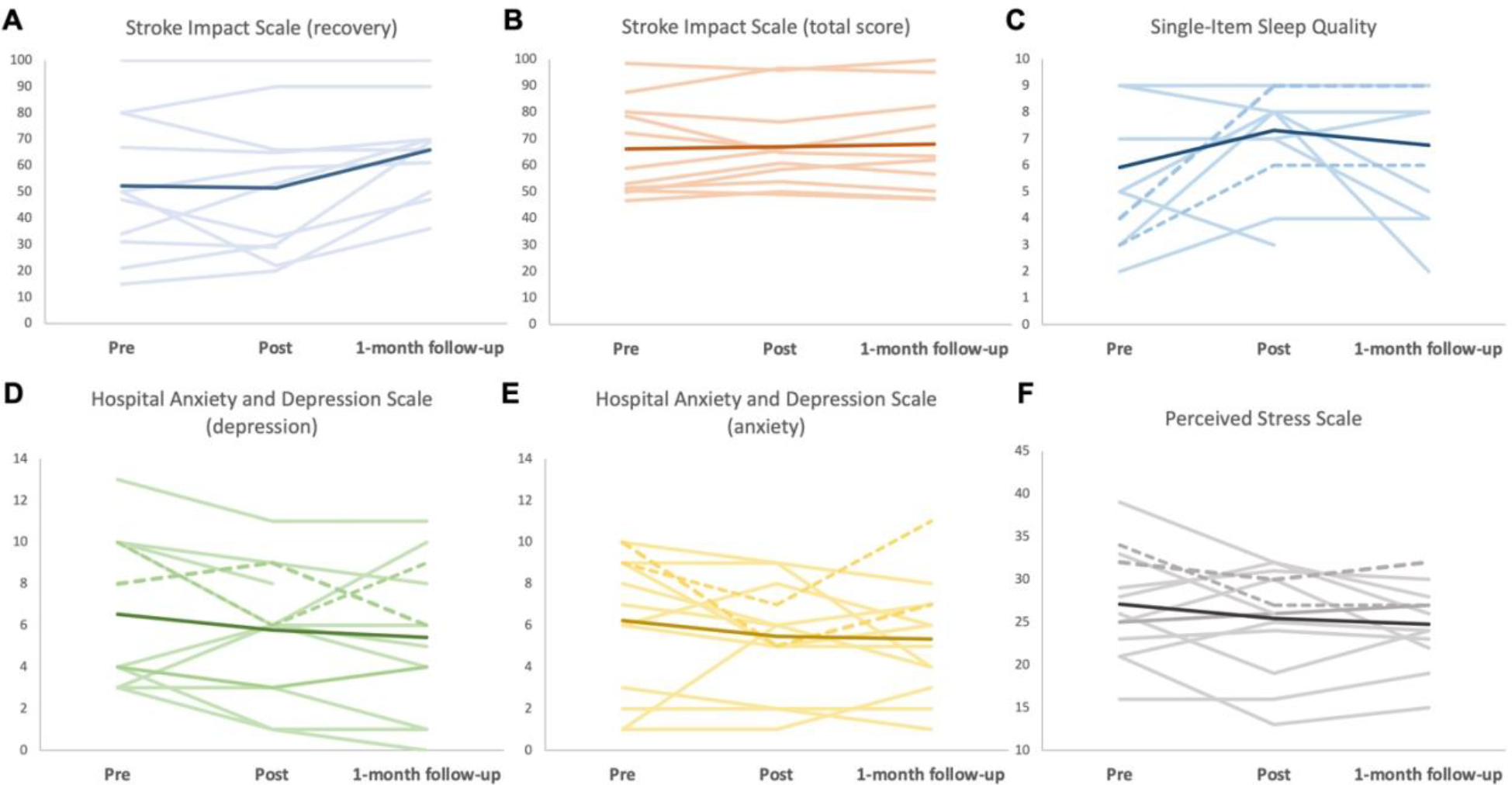
Scores on the quality of life and psychological well-being measures. Individual and mean scores at 3 time points: pre-, post-intervention, and 1-month follow-up. A. Stroke Impact Scale recovery score (0-100%), B. Stroke Impact Scale mean score from all 8 domains (0-100), C. Single-item Sleep Quality score (0-10), D. Hospital Anxiety and Depression score for depression subscale (0-21), E. Hospital Anxiety and Depression score for anxiety subscale (0-21), F. Perceived Stress Scale score (14-70). Thicker lines are mean scores. Thin pale lines are individual scores for stroke survivors (n=11) and dotted lines are individual scores for caregivers (n=2) for C, D, E, and F.

The average usability measured with the PSSUQ was 13.7 ± 10.0 (range from 5-33), which indicates limited usability. Five participants with limited computer savviness had difficulty accessing the website content during the pre-intervention trial. Common difficulties were signing in with their username and password (n=3) and navigating the platform (n=5). Verbal guidance resolved difficulties. Those with greater familiarity with technology experienced no difficulties. One dyad continued to have trouble accessing the content and required support.

## Discussion

This study aimed to evaluate the usability, satisfaction, and potential effectiveness of an online mindfulness intervention for stroke survivors and caregivers. The delivery of an online mindfulness intervention was feasible, with positive overall experiences, high levels of satisfaction, and good adherence to the daily exercises. The qualitative and quantitative data highlighted usability challenges with the online platform. Participants perceived subjective benefits from the intervention, yet no significant changes were observed on self-reported measures. Positive trends from pre to post were noted on the HADS and the SQS. Insights from stroke survivors and participants offered simple strategies to better tailor the intervention, improve the relevance of the exercise, and facilitate accessibility.

Perspectives from a heterogeneous sample of stroke survivors and caregivers were valuable to assess satisfaction, adherence, and relevance of the mindfulness intervention, in the context of a global pandemic. In recent years, more studies explored the benefits of mindfulness-based stress reduction or mindfulness-based cognitive therapy for stroke survivors (Abbott et al., 2014; Lawrence et al., 2013). However, to our knowledge, only two studies captured the experience of stroke survivors with mindfulness (Jani et al., 2018; Wang et al., 2019). Jani et al. (2018) found that a two-hour in-person mindfulness-based stress reduction showed positive acceptability, but participants had trouble maintaining focus and concentration throughout the session. Similarly, Wang et al. (2019) showed that a mindfulness and relaxation program is acceptable, user-friendly, and beneficial for stroke survivors. This is the first study that used Langerian mindfulness with stroke survivors. The integration of mindfulness practice in everyday life seemed particularly appreciated and could be relevant for the clinical applicability and scalability of the intervention. Compared to meditation, Langerian mindfulness with attention to variability requires significantly less time and is easily implementable at a time convenient for the participant, which can vary from day to day.

Many participants experienced difficulties with accessibility and usability of the D2L platform, especially those with limited familiarity with technology. The D2L platform was not perceived as user-friendly, despite its wide use in higher education. The results contrast previous studies that found D2L to be user-friendly and intuitive to use in a tailored academic environment (Alhadreti, 2020; Rucker & Frass, 2017). The primary difference concerns our cohort (older adults, lower levels of education, less exposure to technology and more clinical issues). The results suggest that the initial training and visual guide provided were not sufficient for participants with limited familiarity with technology.

Positive benefits were reported in the semi-structured interview, but this was not observed in the self-reported measures of psychological well-being. The small sample size and large interindividual baseline variability may explain why changes were not detected. Moreover, the evaluation timeline may be too short to observe changes in quality of life and psychological well-being. Participants reported implementing mindfulness strategies in their daily lives, such as changes in mindset during negative events or better ability to remain calm to solve problems in challenging situations. This suggests that the skills acquired during a short three-week online intervention can be transferred to some extent to everyday life. Despite the lack of quantitative changes, the subjective reports seem in line with the results obtained with a similar intervention on people with ALS (Pagnini et al., 2021).

This study is not without limitations. Our sample was small and heterogenous. This study was not powered to detect changes in the self-reported measures, but it provided guidance on the selection of outcome measures for a larger intervention study. The possibility of a social desirability bias is not excluded since participants may have wanted to please the researchers. The results may have limited generalizability due to the small sample size and strong proportion of males and people with right hemisphere stroke. The entire study was conducted during COVID-19, which may have significantly disrupted daily life and afforded time to participate in this kind of intervention.

## Conclusions

To our knowledge, this is the first time a Langerian mindfulness intervention, focused on attention to variability without meditation (e.g., in symptoms, across somatic and cognitive domains), was used with stroke survivors and their caregivers. This study supports the feasibility and acceptance of a three-week online mindfulness intervention for stroke survivors and caregivers. The intervention and clinical assessments were conducted entirely remotely during a global pandemic, which highlight the potential clinical application of such an intervention to allow greater access to individuals with limited mobility or living outside urban centers. This study provided insights about user preferences and accessibility of an online platform designed for higher education. Simplify access to the content, modify exercise wording, offer options to record notes, customize the exercises based on current ability or situation, and provide audio recordings for all modules, are simple suggestions that may help improve the usability of the online platform and the relevance of the intervention. This study is a precursor step in the design of a pilot study and eventually, a potential large, randomized control trial to determine the effectiveness of an online mindfulness intervention with attention to variability for stroke survivors and caregivers.

## Data Availability

Additional quotes from participants are available in the supplementary material.

## Acknowledgements

We thank Sarah Powner for her assistance with portions of data analysis. We acknowledge the early contributions of Didi Matthews, Julie Hershberg, Allison Shapiro and David Karchem for their work on the initial development and modification of the mindfulness intervention. This project was supported by the Southern California Clinical and Translational Science Institute Voucher Program. MD is supported in her position as Postdoctoral Scholar – Visiting Fellow by the Fonds de la Recherche du Québec Santé and the Division of Biokinesiology and Physical Therapy at USC.

## References

Abbott, R. A., Whear, R., Rodgers, L. R., Bethel, A., Thompson Coon, J., Kuyken, W., Stein, K., & Dickens, C. (2014). Effectiveness of mindfulness-based stress reduction and mindfulness based cognitive therapy in vascular disease: A systematic review and meta-analysis of randomised controlled trials. Journal of Psychosomatic Research, 76(5), 341–351. https://doi.org/10.1016/j.jpsychores.2014.02.012

Alexander, C. N., Langer, E. J., Newman, R. I., Chandler, H. M., & Davies, J. L. (1989). Transcendental Meditation, mindfulness, and longevity: An experimental study with the elderly. Journal of Personality and Social Psychology, 57(6), 950–964. http://dx.doi.org.libproxy1.usc.edu/10.1037/0022-3514.57.6.950

Alhadreti, O. (2020). A Comparative Usability Study of Blackboard and Desire2Learn: Students’ Perspective. In P. Zaphiris & A. Ioannou (Eds.), Learning and Collaboration Technologies. Designing, Developing and Deploying Learning Experiences (pp. 3–19). Springer International Publishing. https://doi.org/10.1007/978-3-030-50513-4_1

Ayerbe, L., Ayis, S., Wolfe, C. D. A., & Rudd, A. G. (2013). Natural history, predictors and outcomes of depression after stroke: Systematic review and meta-analysis. The British Journal of Psychiatry, 202(1), 14–21. https://doi.org/10.1192/bjp.bp.111.107664

Braun, V., & Clarke, V. (2006). Using thematic analysis in psychology. Qualitative Research in Psychology, 3(2), 77–101. https://doi.org/10.1191/1478088706qp063oa

Campbell Burton, C. A., Murray, J., Holmes, J., Astin, F., Greenwood, D., & Knapp, P. (2013). Frequency of anxiety after stroke: A systematic review and meta-analysis of observational studies. International Journal of Stroke, 8(7), 545–559. https://doi.org/10.1111/j.1747-4949.2012.00906.x

Colorafi, K. J., & Evans, B. (2016). Qualitative Descriptive Methods in Health Science Research. HERD: Health Environments Research & Design Journal, 9(4), 16–25. https://doi.org/10.1177/1937586715614171

Creswell, J. W., & Plano-Clark, V. L. (2018). Designing and Conducting Mixed Methods Research (3rd ed.). Sage Publications.

Duncan, P. W., Bode, R. K., Lai, S. M., & Perera, S. (2003). Rasch analysis of a new stroke-specific outcome scale: The stroke impact scale. Archives of Physical Medicine and Rehabilitation, 84(7), 950–963. https://doi.org/10.1016/S0003-9993(03)00035-2

Grant, A. M., Langer, E. J., Falk, E., & Capodilupo, C. (2004). Mindful creativity: Drawing to draw distinctions. Creativity Research Journal. https://doi.org/10.1207/s15326934crj1602&3_9

Hartman-Maeir, A., Soroker, N., Ring, H., Avni, N., & Katz, N. (2007). Activities, participation and satisfaction one-year post stroke. Disability and Rehabilitation, 29(7), 559–566. https://doi.org/10.1080/09638280600924996

Jani, B. D., Simpson, R., Lawrence, M., Simpson, S., & Mercer, S. W. (2018). Acceptability of mindfulness from the perspective of stroke survivors and caregivers: A qualitative study. Pilot and Feasibility Studies, 4(1), 57. https://doi.org/10.1186/s40814-018-0244-1

Kupst, M. J., Butt, Z., Stoney, C. M., Griffith, J. W., Salsman, J. M., Folkman, S., & Cella, D. (2015). Assessment of stress and self-efficacy for the NIH Toolbox for Neurological and Behavioral Function. Anxiety, Stress, & Coping, 28(5), 531–544. https://doi.org/10.1080/10615806.2014.994204

Langer, E. J. (2000). Mindful learning. Current Directions in Psychological Science. https://doi.org/10.1111/1467-8721.00099

Langer, E. J. (2009). Counterclockwise: Mindful Health and the Power of Possibility. Ballantine Books.

Langer, E. J. (2012). The mindless use of medical data. Journal of Business Research. https://doi.org/10.1016/j.jbusres.2011.02.018

Lawrence, M., Booth, J., Mercer, S., & Crawford, E. (2013). A Systematic Review of the Benefits of Mindfulness-Based Interventions following Transient Ischemic Attack and Stroke. International Journal of Stroke, 8(6), 465–474. https://doi.org/10.1111/ijs.12135

Lewis, J. R. (1992). Psychometric Evaluation of the Post-Study System Usability Questionnaire: The PSSUQ. Proceedings of the Human Factors Society Annual Meeting, 36(16), 1259–1260. https://doi.org/10.1177/154193129203601617

Mayo, N. E., Wood-Dauphinee, S., Cote, R., Durcan, L., Carlton, J., Côté, R., Durcan, L., & Carlton, J. (2002). Activity, participation, and quality of life 6 months poststroke. Archives of Physical Medicine and Rehabilitation, 83(8), 1035–1042. https://doi.org/10.1053/apmr.2002.33984

Morimoto, T., Schreiner, A. S., & Asano, H. (2003). Caregiver burden and health-related quality of life among Japanese stroke caregivers. Age and Ageing, 32(2), 218–223. https://doi.org/10.1093/ageing/32.2.218

Pagnini, F., Bercovitz, K., & Langer, E. J. (2016). Perceived Control and Mindfulness: Implications for Clinical Practice. Journal of Psychotherapy Integration, 26(2), 91–102. https://doi.org/10.1093/acprof:oso/9780190257040.003.0006

Pagnini, F., & Philips, D. (2015). Being mindful about mindfulness. The Lancet Psychiatry. https://doi.org/10.1016/S2215-0366(15)00041-3

Pagnini, F., Phillips, D., Haulman, A., Bankert, M., Simmons, Z., & Langer, E. (2021). An online non-meditative mindfulness intervention for people with ALS and their caregivers: A randomized controlled trial. Amyotrophic Lateral Sclerosis and Frontotemporal Degeneration, 1–12. https://doi.org/10.1080/21678421.2021.1928707

Patel, M. D., Tilling, K., Lawrence, E., Rudd, A. G., Wolfe, C. D. A., & McKevitt, C. (2006). Relationships between long-term stroke disability, handicap and health-related quality of life. Age and Ageing, 35(3), 273–279. https://doi.org/10.1093/ageing/afj074

Rigby, H., Gubitz, G., & Phillips, S. (2009). A systematic review of caregiver burden following stroke. International Journal of Stroke, 4(4), 285–292. https://doi.org/10.1111/j.1747-4949.2009.00289.x

Shenton, A. K. (2004). Strategies for ensuring trustworthiness in qualitative research projects. Education for Information, 22(2), 63–75. https://doi.org/10.3233/EFI-2004-22201

Snyder, E., Cai, B., DeMuro, C., Morrison, M. F., & Ball, W. (2018). A new single-item sleep quality scale: Results of psychometric evaluation in patients with chronic primary insomnia and depression. Journal of Clinical Sleep Medicine, 14(11), 1849–1857. https://doi.org/10.5664/jcsm.7478

Tickle-Degnen, L. (2013). Nuts and bolts of conducting feasibility studies. American Journal of Occupational Therapy, 67(2), 171–176. https://doi.org/10.5014/ajot.2013.006270

Ulrichsen, K. M., Kaufmann, T., Dørum, E. S., Kolskår, K. K., Richard, G., Alnæs, D., Arneberg, T. J., Westlye, L. T., & Nordvik, J. E. (2016). Clinical Utility of Mindfulness Training in the Treatment of Fatigue After Stroke, Traumatic Brain Injury and Multiple Sclerosis: A Systematic Literature Review and Meta-analysis. Frontiers in Psychology, 7. https://www.frontiersin.org/article/10.3389/fpsyg.2016.00912

Visser-Meily, A., Post, M., Gorter, J. W., Berlekom, S. B. V., Van Den Bos, T., & Lindeman, E. (2006). Rehabilitation of stroke patients needs a family-centred approach. Disability and Rehabilitation, 28(24), 1557–1561. https://doi.org/10.1080/09638280600648215

Wang, X., Smith, C., Ashley, L., & Hyland, M. E. (2019). Tailoring Self-Help Mindfulness and Relaxation Techniques for Stroke Survivors: Examining Preferences, Feasibility and Acceptability. Frontiers in Psychology, 10. https://doi.org/10.3389/fpsyg.2019.00391

World Health Organization. (2004). The World Health Organization Quality of Life (WHOQOL)-BREF. World Health Organization.

Zarit, S. H., Reever, K. E., & Bach-Peterson, J. (1980). Relatives of the impaired elderly: Correlates of feelings of burden. Gerontologist. https://doi.org/10.1093/geront/20.6.649

Zigmond, A. S., & Snaith, R. P. (1983). The Hospital Anxiety and Depression Scale. Acta Psychiatrica Scandinavica, 67(6), 361–370. https://doi.org/10.1111/j.1600-0447.1983.tb09716.x

