## Supplementary material for "Feasibility of an online Langerian mindfulness program for stroke survivors and caregivers"

*Quantitative data collection – self-reported measures*

The Stroke Impact Scale 3.0 (SIS) includes 59 items organized in 8 domains about the impact of stroke on health and life (Duncan et al., 2003). Each question is scored from 1 (severe difficulty) to 5 (no difficulty) and each domain score is transformed. Higher scores indicate lower perceived impact of stroke. Participants also rated their perceived recovery on a sliding scale from 1 (no recovery) to 100 (full recovery). The Hospital Anxiety and Depression Scale (HADS) is a 14-item scale with 7 items each for anxiety and depression subscales (Zigmond & Snaith, 1983). Each item is scored from 0 to 3, with higher scores indicating higher anxiety or depressive symptoms. The Perceived Stress Scale (PSS) includes 10 items about the occurrence of life stressors scored from 0 (never) to 4 (very often) (Kupst et al., 2015). Higher scores indicate higher perceived stress. The Single-item Sleep Quality (SQS) includes an 11-point visual analogue scale about sleep quality in the past week (0: poor, 10: excellent sleep quality) (Snyder et al., 2018). The World Health Organization Quality of Life-BREF (WHOQOL-BREF) comprises 26 questions from 4 domains about health and well-being (World Health Organization, 2004). The scores were transformed, and higher scores indicate better perceived health and well-being. The Zarit Burden Interview includes 22 questions about caregiver burden rated from 0 (never) to 4 (nearly always) (Zarit et al., 1980). Higher scores indicate higher perceived burden. All measures are valid and reliable for stroke survivors or the general population. The adapted Post-Study System Usability Questionnaire (PSSUQ) consists of 5 questions on perceived satisfaction with the website (Lewis, 1992). Each question is scored on a 7-point Likert scale from 1 (Strongly Agree) to 7 (Strongly Disagree). Higher scores indicate lower usability.

*Detailed qualitative data analysis*

Two coders performed inductive thematic analysis to triangulate the data analysis. Specifically, after reading each verbatim multiple times, every meaningful piece of text was assigned to a descriptive label (i.e., code). Each code was entered in a codebook, which listed the codes with a corresponding definition. Individual codes were grouped, and a sub-theme label was applied to capture a pattern. Emerging themes were identified from individual codes and sub-themes.

*Reflexivity and research team*

MD is a female postdoctoral research fellow with >10 years of qualitative and clinical experience with stroke survivors. Her experience allowed her to understand the realities of stroke survivors and caregivers, but it might also have impacted negative responses from participants not wanting to disappoint the interviewer. BC is a female research assistant and at the time was a master’s student in stem cell and regenerative medicine at the USC Keck School of Medicine. She received training about qualitative methods from the research team. Personal assumptions and reflections made during data collection and analysis were noted in a reflective journal and discussed with the larger research team.

**References**

Duncan, P. W., Bode, R. K., Lai, S. M., & Perera, S. (2003). Rasch analysis of a new stroke-specific outcome scale: The stroke impact scale. *Archives of Physical Medicine and Rehabilitation*, *84*(7), 950–963. https://doi.org/10.1016/S0003-9993(03)00035-2

Kupst, M. J., Butt, Z., Stoney, C. M., Griffith, J. W., Salsman, J. M., Folkman, S., & Cella, D. (2015). Assessment of stress and self-efficacy for the NIH Toolbox for Neurological and Behavioral Function. *Anxiety, Stress, & Coping*, *28*(5), 531–544. https://doi.org/10.1080/10615806.2014.994204

Lewis, J. R. (1992). Psychometric Evaluation of the Post-Study System Usability Questionnaire: The PSSUQ. *Proceedings of the Human Factors Society Annual Meeting*, *36*(16), 1259–1260. https://doi.org/10.1177/154193129203601617

Snyder, E., Cai, B., DeMuro, C., Morrison, M. F., & Ball, W. (2018). A new single-item sleep quality scale: Results of psychometric evaluation in patients with chronic primary insomnia and depression. *Journal of Clinical Sleep Medicine*, *14*(11), 1849–1857. https://doi.org/10.5664/jcsm.7478

World Health Organization. (2004). The World Health Organization Quality of Life (WHOQOL)-BREF. *World Health Organization*.

Zarit, S. H., Reever, K. E., & Bach-Peterson, J. (1980). Relatives of the impaired elderly: Correlates of feelings of burden. *Gerontologist*. https://doi.org/10.1093/geront/20.6.649

Zigmond, A. S., & Snaith, R. P. (1983). The Hospital Anxiety and Depression Scale. *Acta Psychiatrica Scandinavica*, *67*(6), 361–370. https://doi.org/10.1111/j.1600-0447.1983.tb09716.x

**Figure S1: Flow chart of study procedure**

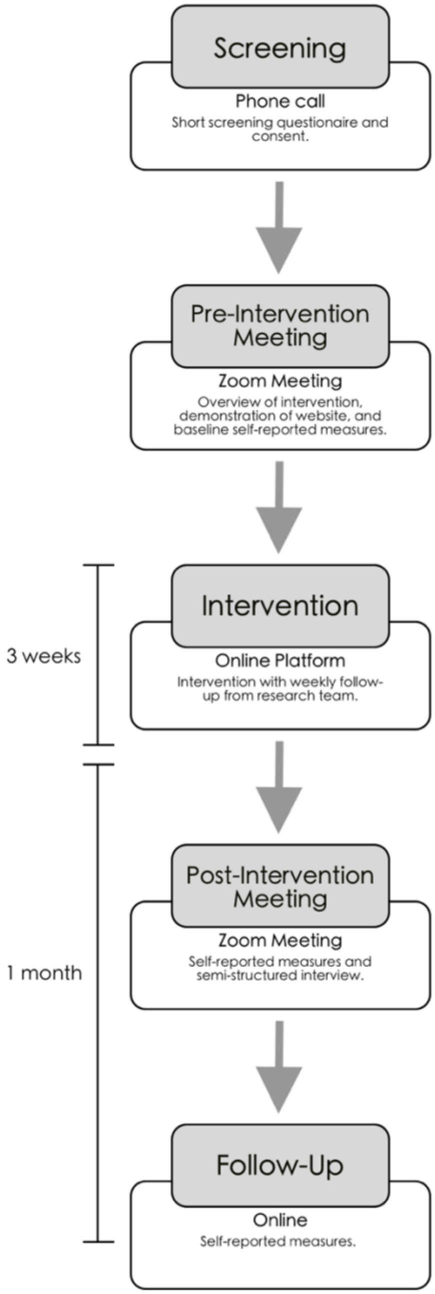

**Figure S2: Concept map of the code, themes and sub-themes that emerged from the thematic content analysis**

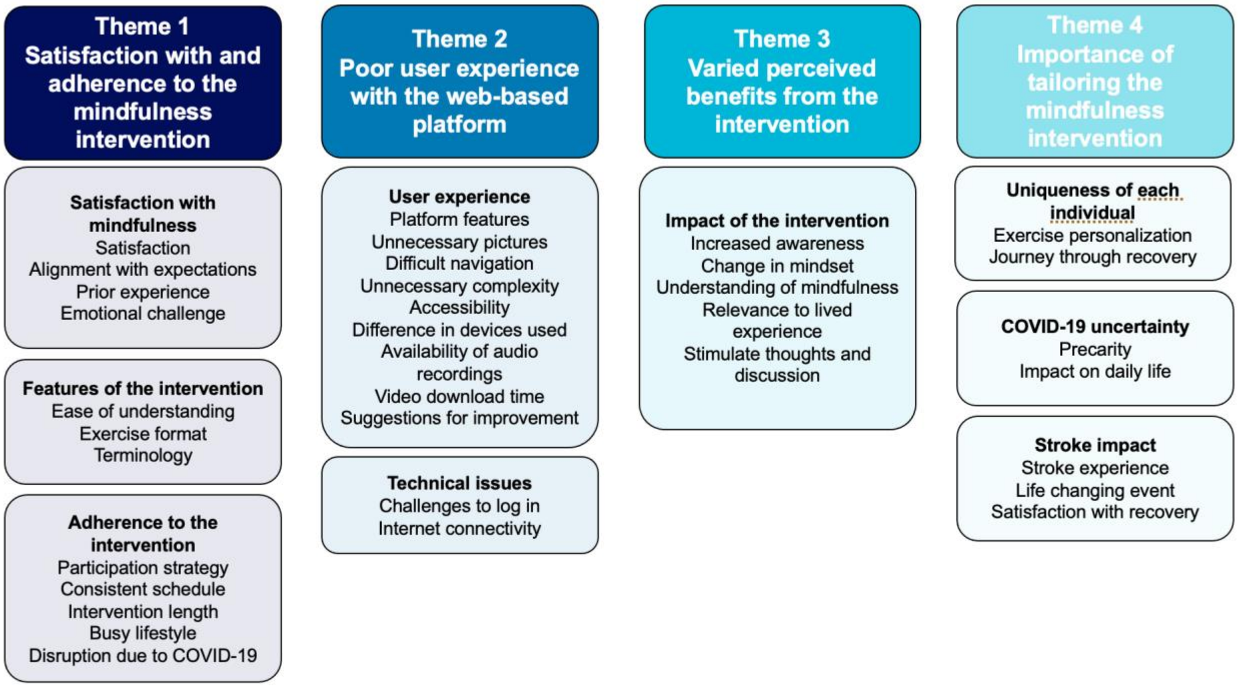

### Interview guide

**Introduction:**
We want to thank you again for your participation in the mindfulness program. Now that you have completed the 3-week program, we want to give you an opportunity to tell us about your experience with the program and to gauge your overall level of satisfaction. We value your opinion and thank you for your willingness to undertake this interview with us.

This discussion is designed to assess your current thoughts and feelings about the program. We also want to learn about the accessibility of the website used to host the program. You should try to convey your honest and true opinion. There is no right or wrong response. Your feedback will help us to make modifications to the program to make sure it is suitable and accessible for [*insert* stroke survivors or caregiver] like you. The discussion will take no more than 30 minutes. May I record the discussion to facilitate its recollection? (if yes, switch on the meeting recording).

**Anonymity:**
Despite being recorded, I would like to assure you that the discussion will be anonymous. The meeting recording will be transferred to a password-protected computer until it is transcribed word for word. Then, the recording will be destroyed.

1. **Overall experience**

What was your opinion about mindfulness in general before starting this study?
Now, tell me about your experience with the mindfulness program you just completed.
What is your overall impression of the mindfulness program?
How satisfied were you with the mindfulness program? Why?

2. **Website accessibility**

*We will now talk about your experience using the Desire2Learn platform.*
Did you access the website on your phone, on a tablet or on the computer?
What did you think of the website?
Please describe any technical problems you experienced during the study.
How accessible was the website for someone who is recovering from a stroke?
How do you think the website could be improved?

3. **Satisfaction with program content**

*We will now talk about your experience with the mindfulness program, itself.*
What did you think of the content of the program?
How align was the program with your expectations?
How clear and easy to understand were the introduction to each module?
How easy to understand were the daily exercises?
How appropriate was the level of language?
What changes can we make to the wording of the exercises to make it easier to understand?
Tell me about how relevant the daily exercises were.
How can the exercises be modified to be more relevant?
How useful were the audio-recordings that went along with the written text?
How useful were the introduction to each module?

How useful were the written description of each exercise?

4. **Adherence to the program**

Please describe what your participation in the program looked like.
How many exercises were you doing on average each day?
How many days per week did you participate in the program?
What made it easy or difficult to follow the daily exercises?
What did you think of the total number of exercises?
What did you think of the duration of the program (3 weeks)?

5. **Perception of effectiveness and changes in health status**

In one of the questionnaires, you indicated being [insert recovery percentage from the SIS] % recovered. How satisfied are you with your recovery?
Do you believe the mindfulness program had benefits for you?
If yes, what were these benefits?
If no, why do you think the program was not beneficial for you?
What are the positive elements you are taking out of this program?
What are the negative elements you are taking out of this program?

**Summary of the discussion**

Let’s summarize some of the key points from our discussion. Is there anything else?
Do you have any questions?

**Conclusion**

Thank you for willingness to participate in this study. This has been a very successful and informative discussion. We want to thank you for giving us a frank and honest assessment of your experience. Your opinions will be a valuable asset to us moving forward. I will be in contact with you in 1 month to complete the follow-up evaluation.

### Additional quotes for each theme

#### Theme 1 – Satisfaction with and adherence to the mindfulness intervention

S03: “My overall opinion of the program is that it was enlightening. It was beneficial. Hum, it was difficult at times, just because the questions would affect me differently than they would affect someone else.”

S07: “Originally, mindfulness was something that that was hocus pocus, because I couldn't do it. But this course gave me a different viewpoint, so now I am a believer.”

S08: “I never thought it would be this good.”

S09: “A lot of [the intervention] seemed like stuff I was already doing or there were examples they were giving me that I couldn't do, so I don't know. […] I didn't feel like I had a big impact on [me].”

S12: “I copied topics from a couple sessions of the program, because there's really great suggestions. [The content of the mindfulness intervention] went on stuff that I read, made me pause and it was good.”

S01: “I found them [the exercises] very, very clear and easy to understand.”

C01: “It was the right length.”

S12: “[The number of daily exercises] was good because I had to stay on top of it. I had to stay engaged, because I had a timeline and [21] days of material to go through. And every day every day I had three exercises to go through.”

S11: “I think the program should be longer. Three weeks, I don't think it's enough to get into mindfulness. But the length of each exercise I thought was good. Ten minutes is a good length of time.”

#### Theme 2: Poor user experience with the web-based platform

S07: “It was a little bit tedious or long [to access the content].”

C03: “I don't know why, but I expected once they both logged in to be able just to reach the content. But clicking through the different tabs, I think was difficult for them and if I didn't do it every day, like, I got mixed up too. So, I was by myself clicking [.] but once I finally got to content, it was easy to navigate. I just wish it was just like an immediate login [.] and like you saw what you had to do for that day.”

C02: "For us, [accessing the content] is very hard."

C01: "We had our phones. That's what [..] our cell phones are old and so that was our main problem. Not you, not what you've done, but what we were able to follow."

S05: "OK well the first thing is the pictures that you have are horrible. They're very depressing, and they are not at all like inspiring you to do it. And particularly the picture of the body parts and the hand, like [.] I [.] could barely look at it [.] like [.] that I did not want to it because of the pictures, but I did it. So I thought the pictures were horrible."

**Theme 3 – Varied perceived benefits from the intervention**

S06: "Hum, it helps you [.] take the negative and switch it to a positive."

S08: "We were able to look to some of things differently."

S01: "When I have awakened in the middle of the night, I've been able to go back to sleep and I think that probably has more to do with this program."

S05: "I made some notes and some of the things they have me look at and I actually shared with some friends, like we haven't had a stroke, and said you know if you were thinking about this what would you think about. And so, we had some discussion on it."

S07: "I enjoyed [the intervention] very much. Maybe it's not noticeable, but it definitely helped to change my attitude to a lot of things, because I think I'm more a negative type person. I see the problems right away, and I rarely see the benefits until it's been pointed out."

S12: "I can't short sell the possibilities of complete recovery with time. I can't, I can't predict anything. […] That's important. […] based on the way I am focusing, my world looks a certain way. It's important."

C01: "It's sort of like a meditation, but active meditation. It helps you get your mind from a negative thought to a positive thought."

**Theme 4- Importance to tailor the mindfulness intervention**

S07: "They were actually very relevant. Actually, a few exercises were a little bit advanced, but I think that because they were, I learned from them."

S03: "If you ask me to move my left hand, I can't do that or walk or something [.] It said walk someplace or do something like that. I can't walk."

S01: "Well obviously, I wish I had recovered more especially on my left side and there have been some other impacts of my stroke over the years, other than the immediate ones. I've definitely had some physical issues that have come up in recent years that are manifestations of the stroke or the impact on my brain that are more noticeable now."

S04: "It is hard to say that COVID didn't influence anything. Every single aspect of life is affected by [COVID-19]."

S05: "I also felt like, because of COVID, it wasn't, like, updated and we didn't verbally update like (.) like the going shopping and stuff like you I can't do any of that because of COVID, so to even imagine that was just even more frustrating and that's not your fault that's just where this landed to be in the process, but I think if you people are going to be doing it, and it's during COVID, you want to adjust the questions. Or just answer as if you were able to go out or something like that."

S10: "So sometimes they would ask me to do things that I wasn't capable of doing. I'm not, for instance, going to the store right now. […] So, would it make more sense if we were to refine some of the exercises to make them more targeted."
